# Incidence of Respiratory distress and its predictors among neonates admitted at neonatal intensive care unit, Black Lion Specialized Hospital, Addis Ababa, Ethiopia, 2018

**DOI:** 10.1101/19007823

**Authors:** Yared Asmare, Hussien Mekonen, Tadesse Yirga, Tesfa Dejenie Habtewold, Aklilu Endalamaw, Wondimeneh Shibabaw

## Abstract

**Background:** Although Respiratory distress is one of the major causes of neonatal morbidity and mortality throughout the globe, it is a serious concern more of in resource limited nations, like Ethiopia. Besides, few studies are available in developing countries. Data from different settings is needed to tackle it. Therefore, we intended to assess the incidence and predictors of respiratory distress among neonates who were admitted in neonatal Intensive care unit (NICU) at Black Lion Specialized Hospital, Ethiopia.

**Methods:** Institution-based retrospective follow-up study was conducted among 571 neonates from January 2013 to March 2018. Data were collected by reviewing patients chart using systematic sampling technique with a pretested checklist; entered using Epi-data 4.2 and analyzed with STATA 14. Median time, Kaplan-Meier survival estimation curve and Log-rank test were computed. Bivariable and multivariable Gompertz parametric hazards models were fitted to detect the determinant of respiratory distress. Hazard ratio with a 95% confidence interval was calculated. Variables with reported p-values < 0.05 were considered statistically significant.

**Results:** The proportion of respiratory distress among of neonates admitted in Black Lion specialized hospital neonatal intensive care unit was 42.9 % (95%CI: 39.3-46.1%) with incidence of 8.1/100(95%CI: 7.3, 8.9)).Being male [AHR=2.4 (95%CI:1.1,3.1)], neonates born via caesarean section [AHR:1,.9((95%CI:1.6,2.3)], home delivery [AHR :2.9 (95%CI:1.5, 5,2)], maternal diabetes mellitus [AHR 2.3(95%CI: 1.4, 3.6)], preterm birth [AHR:2.9(95%CI:1.6, 5.1)] and APGAR score less than 7 [AHR: 3.1 (95%CI:1.8,5.0)] were found to be significant predictors of respiratory distress.

**Conclusions:** The incidence of respiratory distress among neonates was found to be high. Those neonates delivered at home, delivered through caesarean section, preterm neonates, whose APGAR score<7, and born from diabetic mothers were more likely to develop respiratory distress. All concerned bodies should work on preventing RD and give special attention for multifactorial cause of it. Thus; it is indicated to promote health institutional delivery more. Besides, a need to establish and/or strengthen strategies to prevent the occurrence of respiratory distress among babies with low APGAR score, preterm babies, born from diabetes mellitus mothers, and delivered through caesarean section.

## Background

Respiratory distress (RD) is a common newborn problem immediately following birth. It is typically triggered by abnormal respiratory function during the transition from fetal to neonatal life. Tachypnea, intercostal retraction, nasal flaring, audible grunting, and cyanosis are the common manifestation of RD. The successful transition from fetal to neonatal life requires a series of rapid physiologic changes of the cardiorespiratory system. These changes result in redirection of gas exchange from the placenta to the lung and comprise replacement of alveolar fluid with air and onset of regular breathing [1]. It may be transient; yet, persistent distress requires a rational diagnostic and therapeutic approach to optimize outcome and minimize morbidity.

RD one of the most common reasons for neonates to be admitted to the neonatal intensive care unit (NICU) [2, 3]. Fifteen percent of term infants and 29% of late preterm infants admitted to the NICU develop significant respiratory morbidity. This is even higher for infants born before 34 weeks’ gestation [4].

Certain risk factors increase the likelihood of neonatal RD. Prematurity, low first and fifth minute APGAR score, meconium-stained amniotic fluid (MSAF), caesarian section delivery, gestational diabetes, maternal chorioamnionitis, premature rupture of membranes mother it [5]and oligohydramnios and structural lung abnormalities are some predictors identified with previous studies [4, 6-9]. Other common causes also include transient tachypnea of the newborn, meconium aspiration syndrome, pneumonia, sepsis, pneumothorax, persistent pulmonary hypertension of the newborn [10].In contrast, the risk decreases with each advancing week of gestation and born with spontaneous vaginal delivery [11].

Regardless of the cause, if not recognized and managed quickly, respiratory distress can escalate to respiratory failure and cardiopulmonary arrest and finally leads to death. Therefore, it is imperative that any health care practitioner caring for newborn infants can readily recognize the signs and symptoms of respiratory distress, differentiate various causes, and initiate management strategies to prevent significant complications or death. Consequently, neonates in need of critical medical attention are admitted to the NICU ward. These infants tend to be preterm, low birth weight, or have serious medical conditions including RD [12, 13].

Globally, there are different policies, strategies, and programs which work on prevention and care of preterm birth and its birth outcome including RD, like Sustainable Development Goals (SDGs) and Every Women and Every Child initiative [14, 15]. Despite of this, it is among the leading cause of neonatal mortality and morbidity [16-20].In Ethiopia, according to a report of FMOH, RD is most common cause of neonatal mortality and morbidity [16-21]. So that, it remains to be a major community health problem by increasing the regular price of healthcare for neonates with in the first 28 day of life for unindustrialized nation, including Ethiopia. These great medical expenditures might burden both the parents, families and the community at large. Hence, this is the double agenda to address the survival gap of neonates which needs an inclusive investigation strategy to end the preventable cause deaths of newborns. Additionally, few studies in developing countries, including Ethiopia have provided data needed to tackle it. Therefore, we aimed to determine the incidence and predictors of RD among neonates who were admitted in NICU at Black Lion Specialized Hospital, Ethiopia.

## Methods

### Study design, setting, and population

An institution-based retrospective follow-up study was conducted among a cohort of neonates in the previous consecutive five years. Our source population was all neonates who were admitted in NICU of Black Lion Referral Hospital. All neonates who were admitted to NICU in the previous five consecutive years were considered as the study population.

### Eligibility criteria

All neonates’ medical cards documented in the previous five years from the study period were recruited and incomplete cards were excluded.

#### Sample size determination and sampling procedure

The sample size was determined by using double population proportion formula using Epi-Info version7 by assuming one to one ratio of exposed to non-exposed, 95% level of confidence and power of 80%. We considered four significantly associated factors to calculate the sample size; the larger sample size was 522. After adding 10% non-response rate, the total sample size became 604. The neonate’s card was accessed by using systematic random sampling technique after determining the sampling fraction (k=6) and the first card was selected by lottery method.

### Study variables

#### Dependent variable

Incidence of RDS

### Independent variables

#### Socio-demographic factors

Neonatal-related variables, including age at admission, gestational age, sex, the weight of neonate, Date of NICU admission and discharge. Maternal-related variables were age and residency.

#### Gynecologic-obstetric related factors

having ANC follow up, gravidity, parity, mode of delivery, multiple pregnancies, PROM, preeclampsia, abruption placenta, and breastfeeding initiation

#### Medical disorders in mother

Hypertension, DM, HIV/AIDS

#### Neonatal outcome condition

Apgar score, RD, sepsis, jaundice, hypothermia, PNA, hypoglycemia, meningitis, esophageal atresia, CHD.

#### Data collection tools

Pretested checklist was used to collect the required data from the patient’s chart. The checklist was translated to local language Amharic and back to English; consistency was checked. Data were collected by reviewing the complete patients’ card in the previous five consecutive years from the study period. RD was confirmed by reviewing the medical charts.

#### Data quality control

Data quality was assured by designing proper data abstraction tools. The checklist was evaluated by experienced researchers. The data collection instrument was pre-tested on 5% of the sample size. Rigorous training was given regarding the data abstraction checklist and data collection process for both data collectors and supervisors. During the data collection time, close supervision and monitoring were carried out by supervisors and investigator. Double data entry was also done using Epi data 4.2.0 software.

#### Data processing, analysis, and presentation

Before analysis, data was cleaned, edited and coded. Any errors identified at this time were corrected after review of the original data using the code numbers. Data were entered using Epi-Data version 4.2.0 and analyzed using STATA 14 statistical software. Incidence density rate (IDR) was calculated for the entire study period. Subsequently, the number of RD within the follow-up period was divided by the total person-time at risk on follow-up and reported per 100-person day. Kaplan-Meir was used to estimate median mean survival time and log-rank tests were used to compare survival curves. Proportional hazard assumption was tested both using graphically. We also done the Schoenfeld residual test for all predictors and the global test revealed that the proportional hazard assumption is met. After checking this assumption, by comparing models, a more effective hazard model was selected by using log likelihood ratio (LR) test and the Akaike Information Criterion (AIC). In the parametric approach, the baseline hazard and the vector of its parameters is assessed together with the regression coefficients. The best-fitted model was chosen using AIC; those having the smallest AIC. Then, parametric models were completed for neonates to ascertain the possible predictors. Variables having a p-value less than or equal to 0.05 in the bivariate analysis were fitted to the multivariable Gompertz hazard distribution regression model with a 95% confidence interval. A p-value less than 0.05 were considered as statistically significant.

### Operational definition

#### Event (neonatal respiratory distress)

The presence of two or more of the following signs: an abnormal respiratory rate (tachypnea > 60breaths/min, bradypnea < 30 breaths/minute, respiratory pauses, or apnea) or signs of labored breathing (expiratory grunting, nasal flaring, intercostal recessions, xyphoid recessions), with or without cyanosis.

#### RD

was diagnosed based on the presence of two or more of the following signs: an abnormal respiratory rate, expiratory Grunting, nasal flaring, chest wall recessions and cyanosis, in their files

## Results

### Characteristics of neonates

Among 604 neonates’ charts reviewed, 571 (94.5%) records were met enrollment criteria in the final analysis. Of which, about 299 (52.34%) of the study participants were males. Neonates in late neonatal period account more than half of the study participant. The mean age of the cohort at the time of admission to NICU was 3 ± 3.72 SD days. In this finding, more than half of neonates admitted to NICU were diagnosed with neonatal sepsis. The other common causes of admissions were jaundice, hypothermia and PNA. In addition, the common types RD for neonatal admission was RDS or hyaline member diseases (See figure 1)

**Figure 1:**
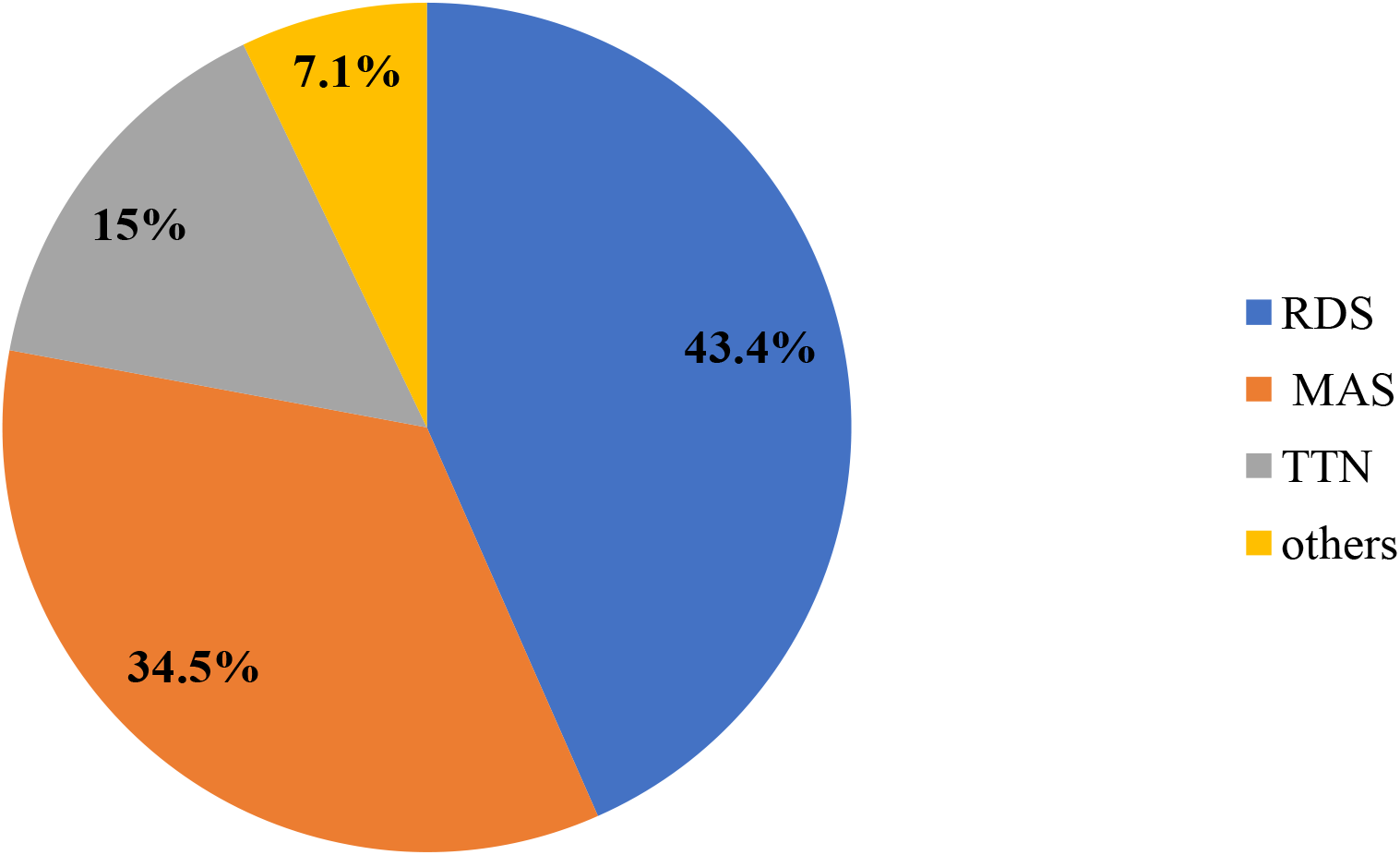
common types of RD for neonates admitted in NICU at Black Lion Specialized Hospital, 2018, (n = 571).

**Table 1:**
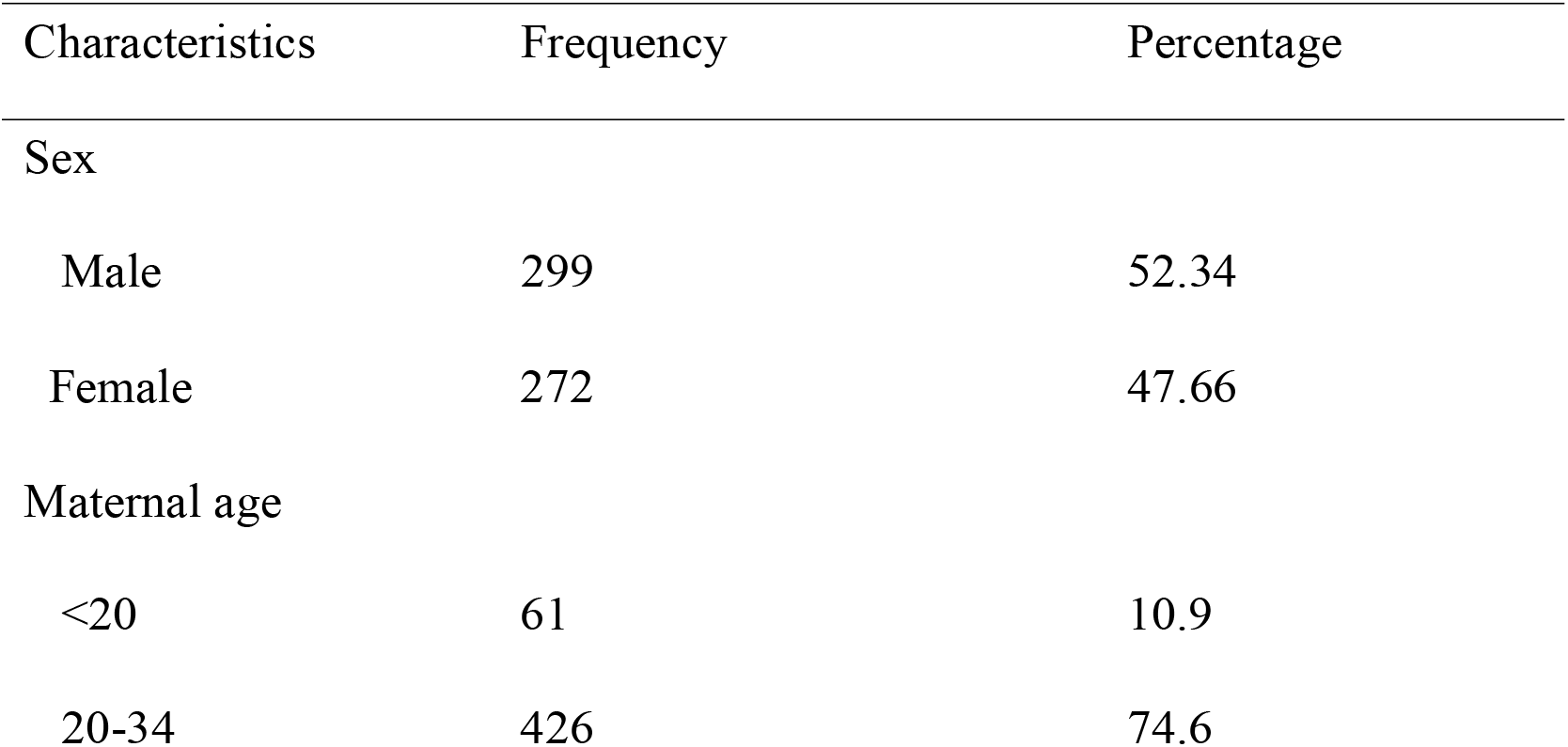
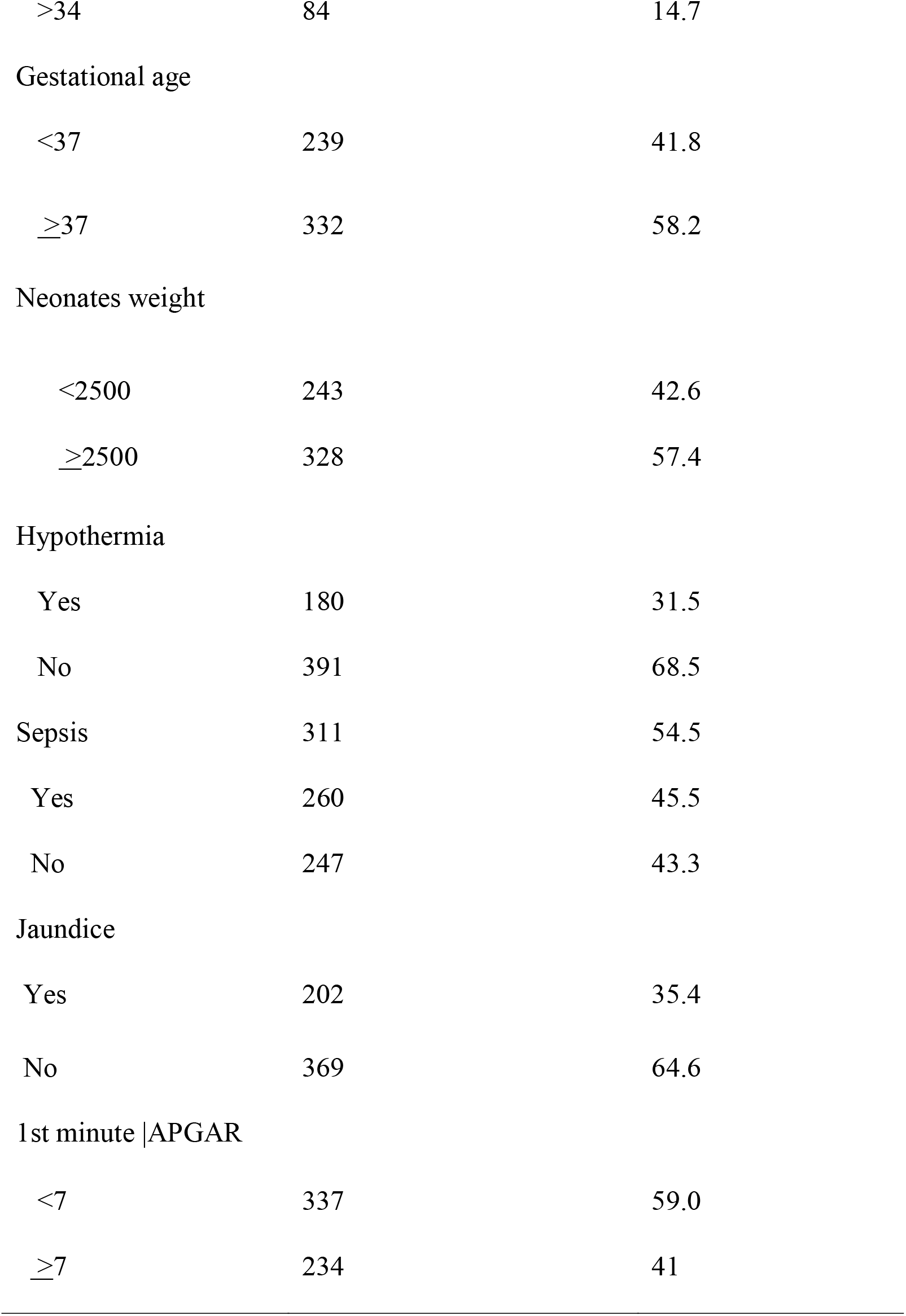
Characteristics of neonates admitted in NICU at Black Lion specialized Hospital, 2018 (n = 571).

### Socio-demographic and obstetric characteristics of mothers

In the current study, most of the mothers were found between the ages of 20-34. The mean age of mothers was found to be 28±5.42SD years old. Among the total mothers enrolled into the study, 336(58.9%) of the mothers had given a birth through spontaneous vaginal delivery and half of neonate that was delivered via CS was developed RD during the follow-up period. Regarding obstetric, gynecological and medical diagnose of maternal diseases, PROM 250(43.8%), HIV/ADIS and DM were identified. The results of this study also indicated that majority of 402(70.4%) neonates of their mother had ANC follow-up. (See table two).

**Table 2:**
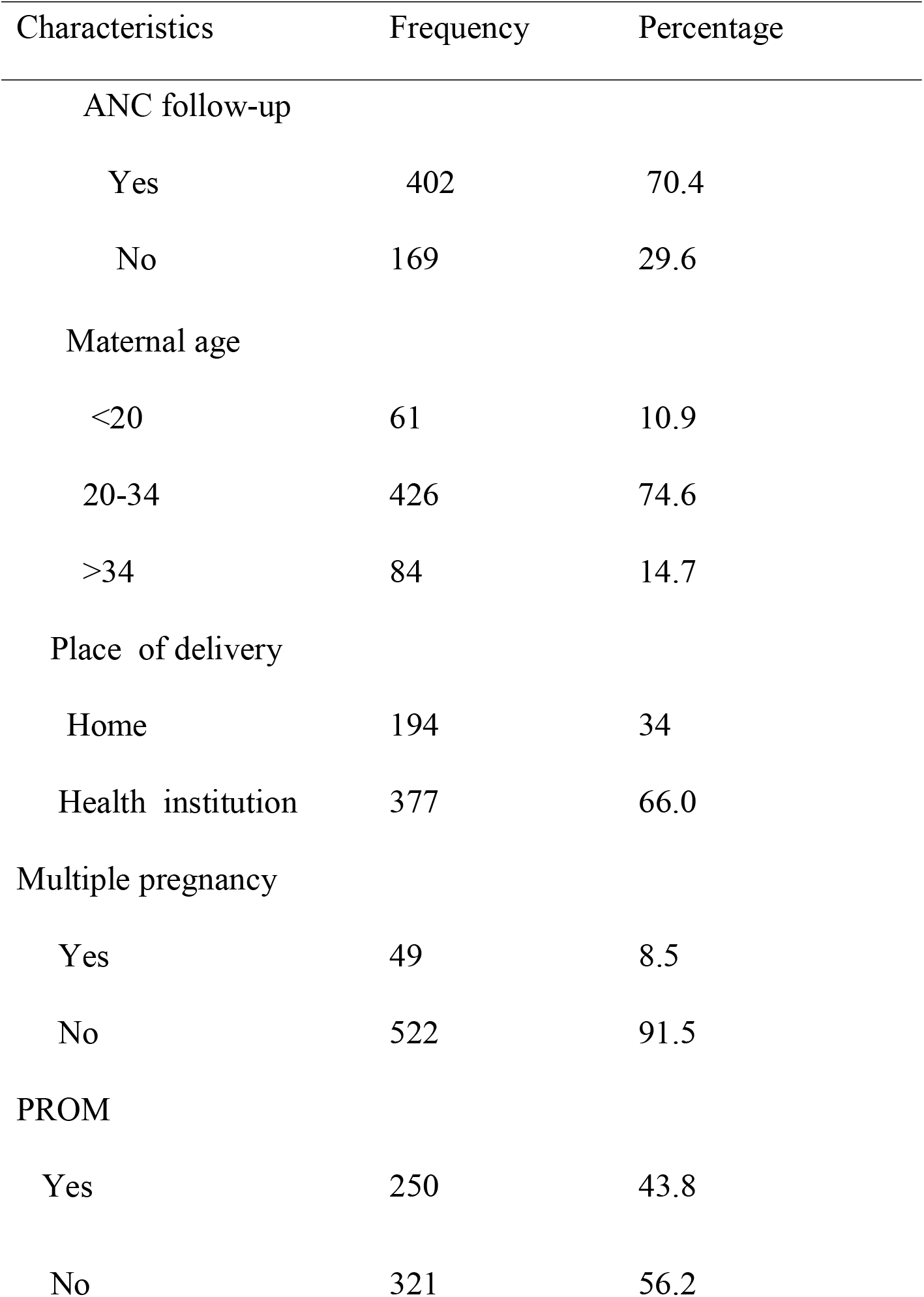
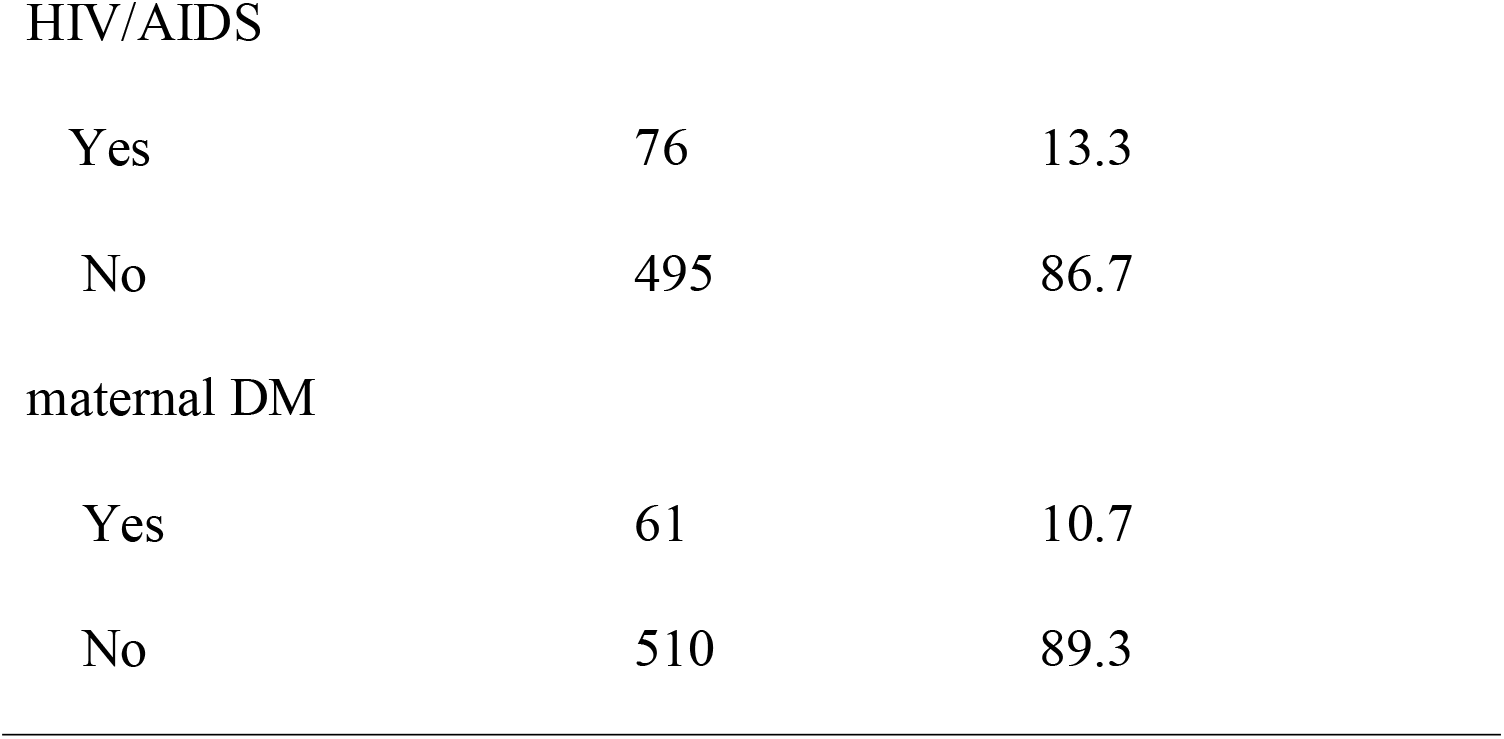
Socio-demographic and obstetric characteristics of mothers of neonates admitted in NICU at Black Lion specialized Hospital from January 2018, (n = 571).

| Characteristics | Frequency | Percentage |
| --- | --- | --- |
| ANC follow-up |  |  |
| Yes | 402 | 70.4 |
| No | 169 | 29.6 |
| Maternal age |  |  |
| <20 | 61 | 10.9 |
| 20-34 | 426 | 74.6 |
| >34 | 84 | 14.7 |
| Place of delivery |  |  |
| Home | 194 | 34 |
| Health institution | 377 | 66.0 |
| Multiple pregnancy |  |  |
| Yes | 49 | 8.5 |
| No | 522 | 91.5 |
| PROM |  |  |
| Yes | 250 | 43.8 |
| No | 321 | 56.2 |
| HIV/AIDS |  |  |
| Yes | 76 | 13.3 |
| No | 495 | 86.7 |
| maternal DM |  |  |
| Yes | 61 | 10.7 |
| No | 510 | 89.3 |

### Overall Proportion and incidence rate of RD in neonates

This finding showed that 245(42.9%) with (95%CI: 39.3-46.1)) were developed RD. The overall incidence of RD was also found to be 8.1 per 100 neonate day with 95%CI :(7.29, 8.9) with 4331-person day observation.

### Time to discharge of neonates with RD

The overall median length of hospital stay for neonates with RD under the study was 9 days with (95%CI; 8-10) and overall length of hospital stay were 28 neonates days with an interquartile range of (5, 30) neonate-days. The cumulative probability of neonates not to be developed RD at the end of the first day was 94.4%, at fifth to 10^th^ days was 41.3%, and at 20–28days was 19.14% (See figure 2).

**Figure two:**
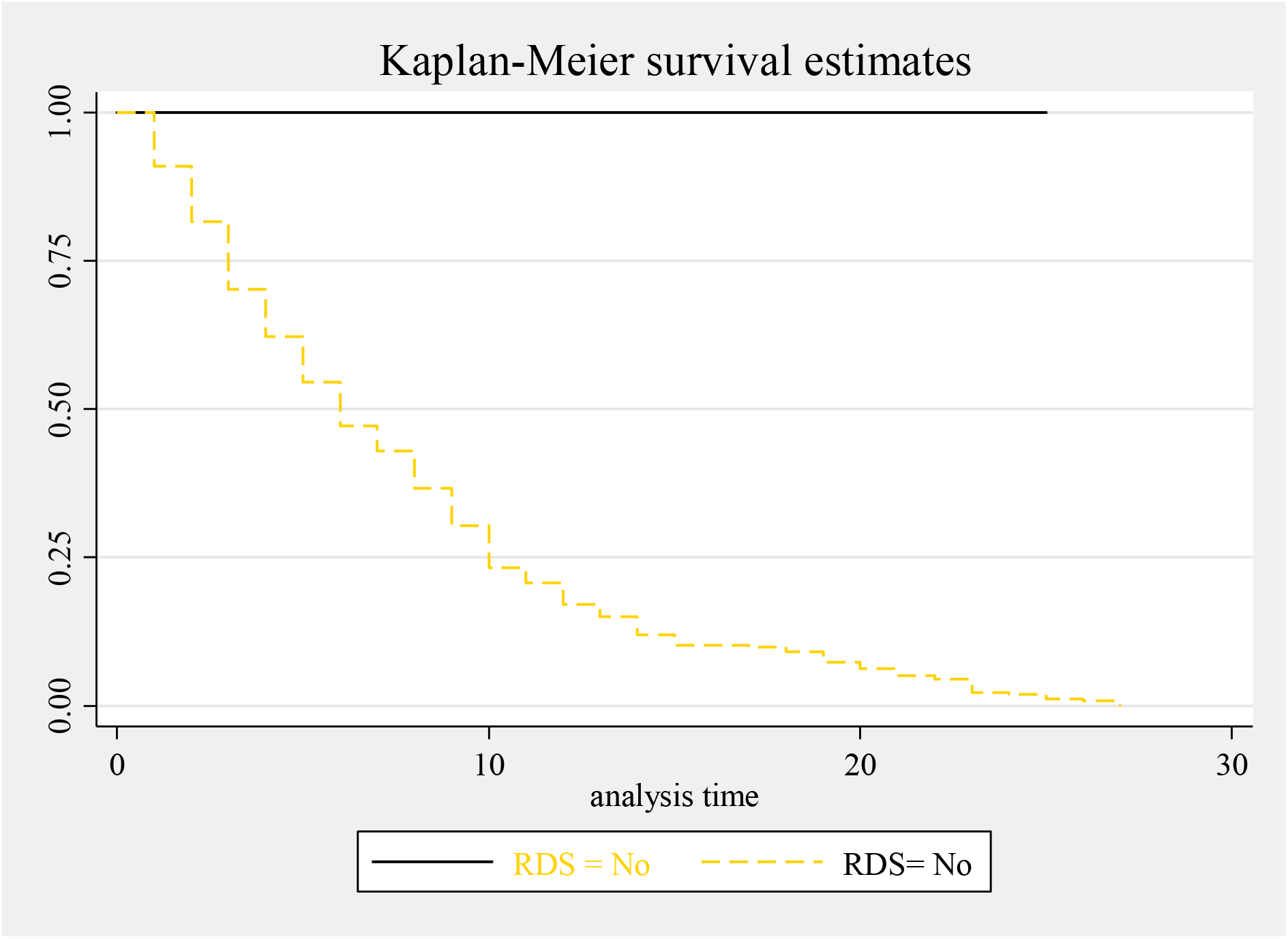
Overall Kaplan-Meier survival estimate of neonates with RD admitted in NICU. Proportional hazard assumption was evaluated using Kaplan Meier survival and Sheffield residual global test and it was met (x^2^ = 5.11 = P-Value =0.08)(See figure three).

### Model comparison criteria

The goodness of model fitness was checked using the Cox-Snell residual test. Based on the Akaike Information Criterion, the univariate Gompertz hazard distribution (AIC = 435.8) model was more efficient than parametric exponential (AIC = 987.5) and Weibull (AIC = 686.9) semi parametric Cox-proportional hazard (AIC =1123.54) model (See figure 4).

**Figure 3:**
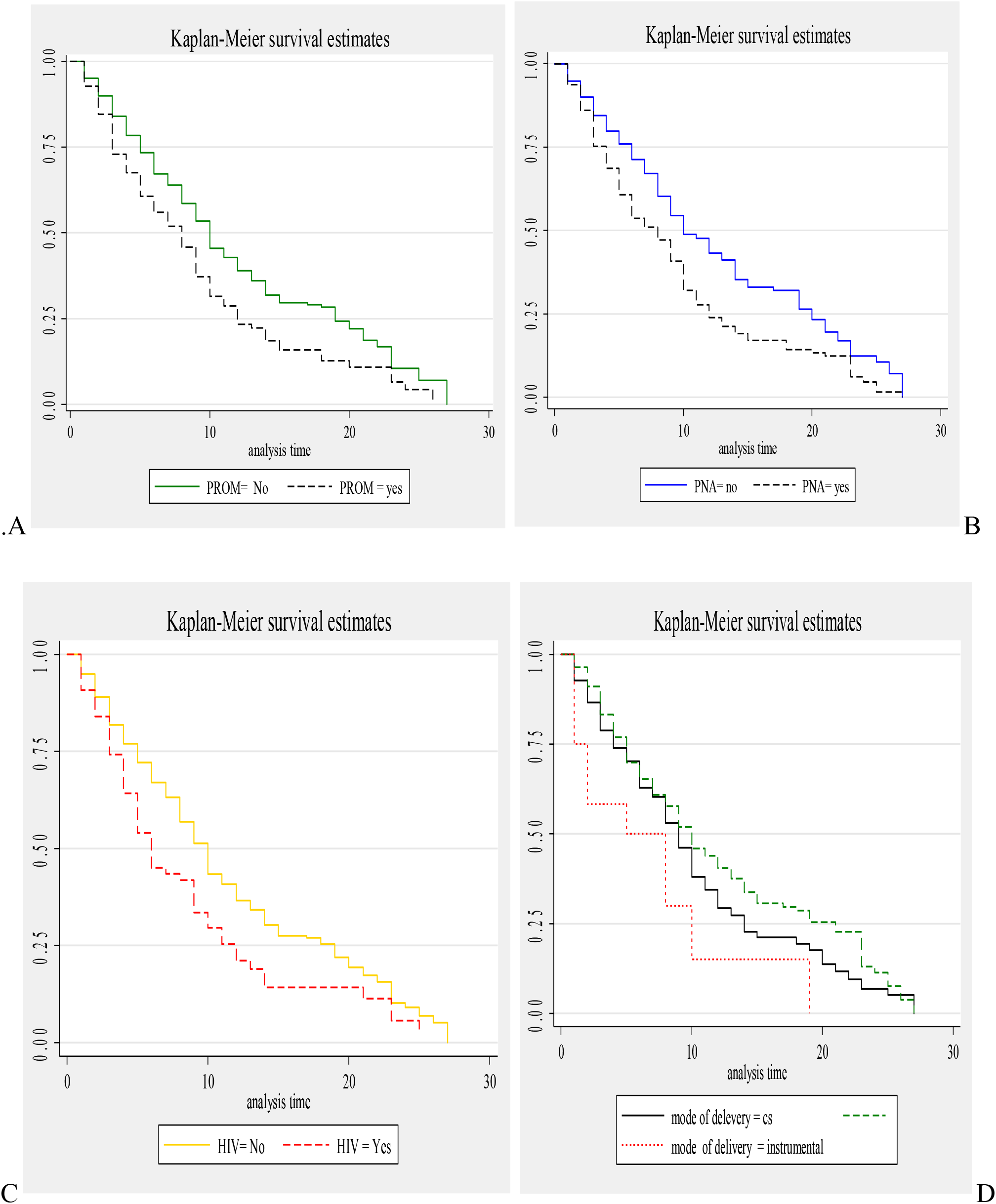
Kaplan-Meier survival curve of neonates with RDS admitted in NICU by **A**:PROM, **B**:PNA, **C**:having maternal HIV/ADIS and **D**:mode of delivery.

**Figure 4:**
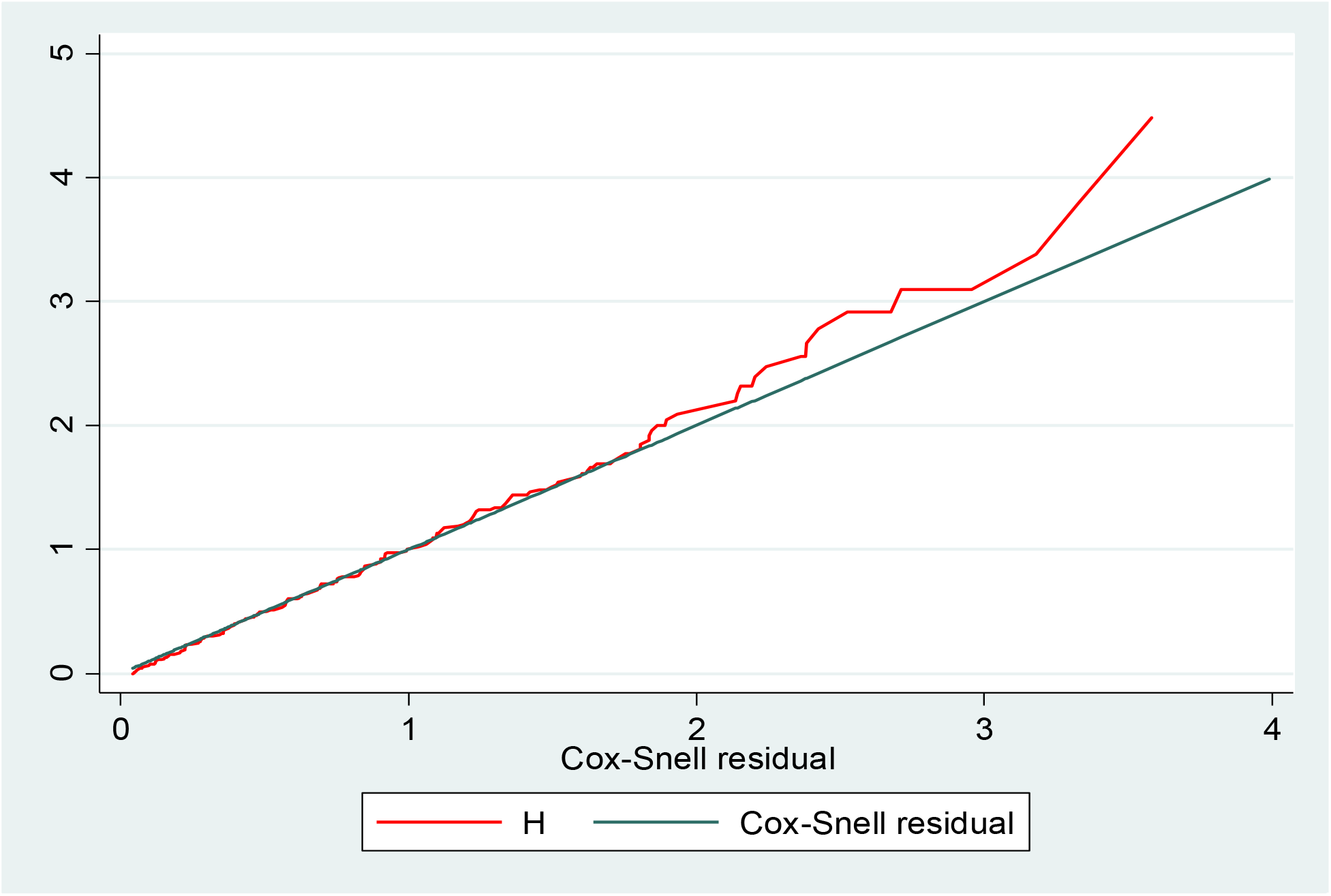
Cox-Snell residual Nelson-Alen cumulative hazard graph on neonates with RDS admitted in NICU at Black LionSpecialized hospital, Ethiopia, 2018 (n = 571).

### Predictors of respiratory distress

The univariate and multivariable parametric Gompertz hazard distribution regression model was used to identify predictors of RD for neonates from admission to discharge in the neonatal intensive care unit. Findings from bivariate analysis showed that weight for gestational, being male, having no antenatal follow-up, multiple pregnancy, neonates born via caesarean section, home delivery, PROM, maternal DM and HIV/ADIS, preterm birth, neonatal sepsis and APGAR score less than 7 were significantly associated with time to discharge of neonates with RD. However, in the multi-variable analysis being male, neonates born via caesarean section, home delivery, maternal DM, preterm birth, neonatal sepsis, PROM and Apgar score less than 7 were continued statistically significant predictors of RD. The hazard of RD in male neonates was 2.4 times increase than their counterpart [AHR: 2.4(95%CI: 1.1, 3.1)] The current study also showed that the hazard of RD among neonates born via caesarean section was nearly 2 times increased hazard than neonates born through vaginally [AHR:1,.9((95%CI:1.6,2.3)]. In this study, the risk of RD for neonates born at home was almost 3 times higher than those delivered at health institution [AHR :2.9 (95%CI:1.5, 5,2)]. This result also indicated that neonates delivered from mothers who had DM increased the risk of RD by 2.3 times as compared with their counterpart [AHR 2.3(95%CI: 1.4, 3.6)]. As the gestational age increases in 1 week the rate of RD s was decreased by 10% [AHR: 2.9(95%CI: 1.6, 5.1)]. The risk of RD was also increased by three times for a neonate who had APGAR score less than 7 as compared with neonates having APGAR score greater than or equal to 7[AHR: 3.1 (95%CI:1.8,5.0)]. Additionally, neonatal sepsis increases the risk of RD BY 60% [AHR :1.6(95%CI:1.1, 2.4)]. The last predictor for RD was born from mothers with PROM. So that neonates born from PROM had 1.1 times higher risk of RD than their counter parts [AHR: 1.1(95%CI: 1.8, 1.5)].

**Table 3:**
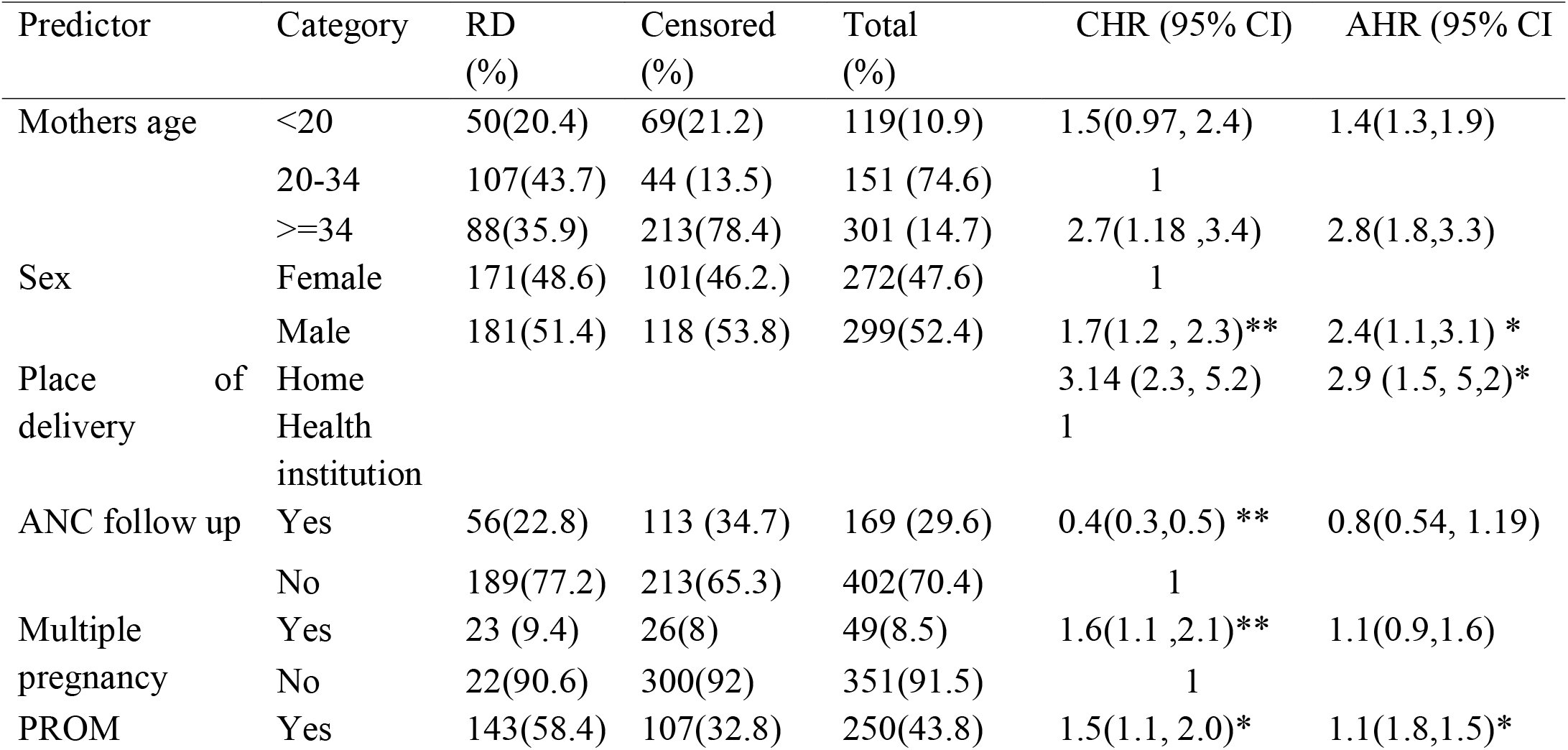
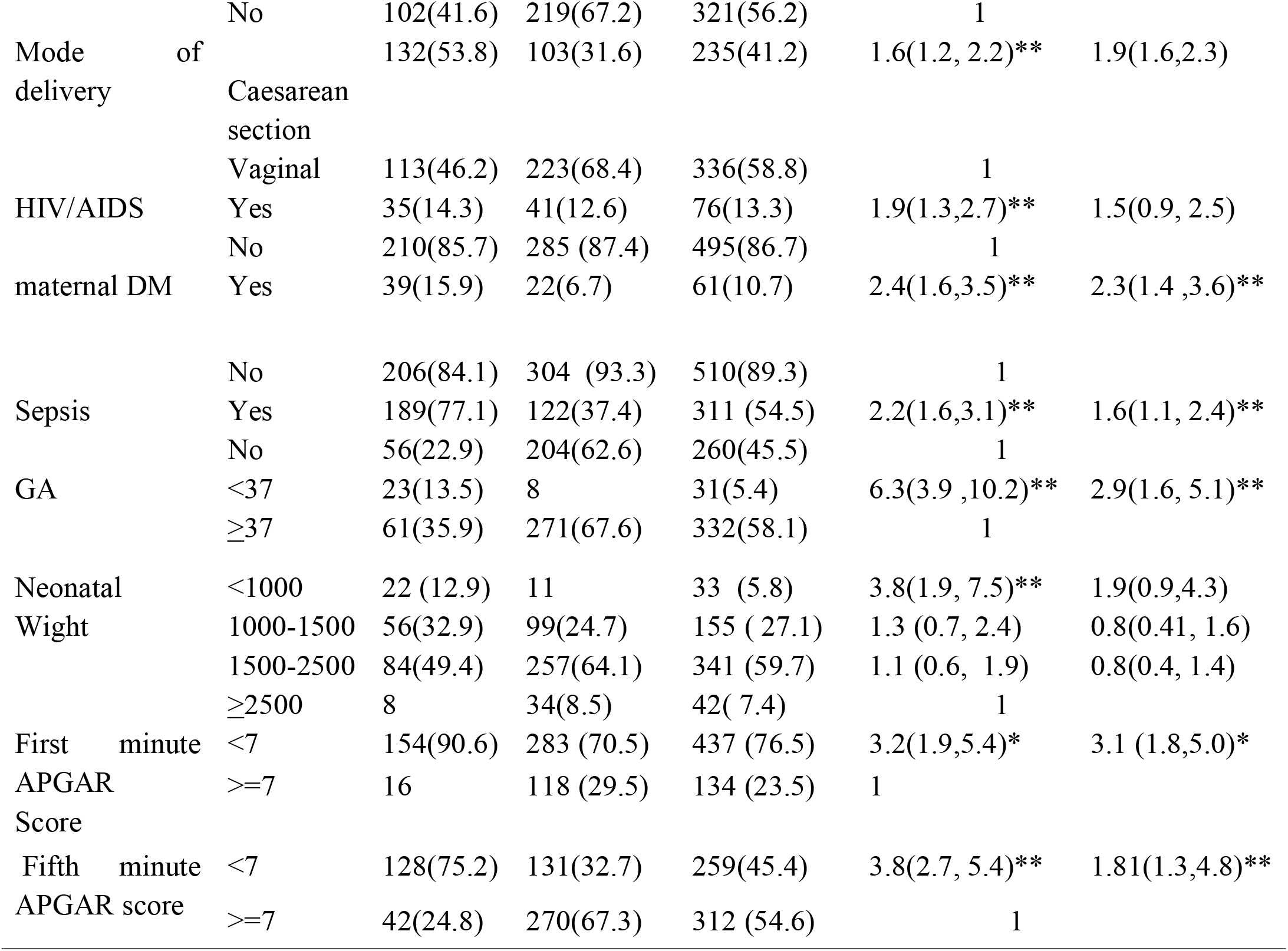
Gompertz hazard model for predictors of incidence of RD among neonates admitted in NICU at Black Lion Specialized referral Hospital, 2018 (n = 571).

## Discussion

In the current study, the overall proportion of RD among neonates admitted in Black lion specialized hospital neonatal intensive care unit was 42.9 %(95%CI: 39.3-46.1%). This finding is in line with a study reported in Republic of China [22]. This might be due to the study done in china is a meta-analysis which includes 26 studies which assess the association between CS and the risk of neonatal RDS, which might increase the proportion of RD.

However, our finding is higher than a study conducted in several countries reported by Nepal Medical College and Teaching Hospital 34% [23], India 33.4% [9], Egypt 34.3 [24], National Institute of Child Health, Karachi 4.6% [25], Northern Italian 20.1% [26] and Portugal (8.83%) [27].

This might be due to the difference in the study setting, in which most of the study that we used to compare was from developed countries in which neonatal, maternal health care service might have an advanced and it will reduce the proportion of neonatal birth with RD.

In contrast, the finding is lower than a report in Saudi Arabia 54.7%[7],Cameroon 47.5%[6] and Poland 54,29%[28]. This discrepancy might be due to difference in sample size and characteristics of the study participants. A study in Poland for example includes a small sample size and they try to assess the incidence of only respiratory distress syndrome which might increase the incidence of RD.

Based on the current finding, the overall incidence of RD was 8.1 (95%CI: 7.3, 8.9) per 100 neonate-date. The common causes of respiratory distress in our study were respiratory distress syndrome and meconium aspiration. This is true in the finding from Nepal and Egypt [23, 24].

The predictor of RD were not a single problem\ Preterm birth, caesarean section delivery, APGAR score<7, sepsis, PROM and maternal diabetes mellitus and home delivery were found to be predictors of RD. The risk of RD in male neonates was 2.4 times increase in than their counterparts. This finding is similar with a study done in China [27] and Cameroon [6]. This finding is supported by the scientific evidence that male neonates have been reported to have a higher level of circulating testosterone than females which might be associated with differences in pulmonary biomechanics and vascular development that lead to increased respiratory related morbidity among males.[29].This finding also showed that the risk of RD among neonates born via caesarean section was nearly 2 times increased hazard than neonates born through vaginally. This finding was in line with the results reported in china [22], Cameroon [6] and Italy [30].

In this study, the risk of RD for neonates born at home was almost 3 times higher than those delivered at health institution. This result also indicated that neonates delivered from mothers who had DM increased the risk of RD by 2.3 times as compared with their counterpart. This finding was supported with a study done in China [31].This might be neonates born from DM women may have plentiful glucose stores in, but develops hypoglycemia because of high insulin secretion induced by maternal and fetal hyperglycemia. This might leads to increase metabolic demand which leads to increase in respiratory efforts.

Similarly, this study presented that a neonate who was preterm at birth was nearly 3 times at greater hazard of having RD compared to those who were term birth. The current finding is also supported by a study done in Cameron [6] and Italy [30]. This was supported by the proven evidence that as gestational age increases fetal development will be increase and hazard of developing different fatal complications including RD related with prematurity could decline and risk of RD will be reduced as a result of lung maturity and increase production of surfactants[6]. The risk of RD was also increased by three times for a neonate who had APGAR score less than 7 as compared with neonates having APGAR score greater than or equal to 7.This finding is also reported by the following studies[6] [25] [26]. Neonatal sepsis was significantly associated with the risk of developing RD. This finding is also similar with a study done in Nepal and Egypt, [23, 24].

## Conclusion

The incidence of respiratory distress among neonates was found to be high. Those neonates delivered at home, delivered through caesarean section, preterm neonates, whose APGAR score<7, and born from diabetic mothers were more likely to develop respiratory distress. Thus, it is indicated to promote health institutional delivery more. Besides, a need to establish and/or strengthen strategies to prevent the occurrence of respiratory distress among babies with low APGAR score, preterm babies, born from diabetes mellitus mothers, and delivered through caesarean

## Data Availability

The data used in this manuscript can be easily availed with email contact of the main authors. Data made available to all interested researchers upon request and the supporting data of the conclusions of the study is available at the abstract.

## Abbreviations

APGAR: Appearance pulse grimace activity respiration
CI: Confidence interval
COR: Crude odd ratio
GA: Gestational age
HIV: Human immunodeficiency virus
HMD: Hyaline membrane disease
HR: Hazard ratio
NICU: Neonatal intensive care unit
PNA: Perinatal asphyxia
PROM: Prolonged rupture of membrane
RD: Respiratory distress.

## Declarations

### Ethical consideration

Ethical clearance was obtained from AAU, college of nursing and midwifery research committee. Then Letters of cooperation was written to TASH and concerned bodies. Permission was obtained from the clinical director and subsequent department and unit heads of the hospital. Following these, searching and obtaining of the selected samples’ medical record was processed with the assigned person. Finally, Care has been taken from disclosing patient’s records.

### Consent for publication

The study was no a clinical trial that needs direct involvement of the patient and not elicit an injury to participant since the data is collected from the patient chart. so that no any need of consent for publication.

### Author’s contribution

All authors contributed to the design of the study and the interpretation of data. YA performed the data analysis and compiled the whole work. YA and HM drafted the manuscript. All authors critically revised the draft manuscript. All authors read and approved the final manuscript.

